# Access to personal protective equipment in healthcare workers during the COVID-19 pandemic in the United Kingdom: results from a nationwide cohort study (UK-REACH)

**DOI:** 10.1101/2021.09.16.21263629

**Authors:** Christopher A. Martin, Daniel Pan, Joshua Nazareth, Avinash Aujayeb, Luke Bryant, Sue Carr, Laura J. Gray, Bindu Gregary, Amit Gupta, Anna L. Guyatt, Alan Gopal, Thomas Hine, Catherine John, I Chris McManus, Carl Melbourne, Laura B. Nellums, Rubina Reza, Sandra Simpson, Martin D. Tobin, Katherine Woolf, Stephen Zingwe, Kamlesh Khunti, Manish Pareek, On behalf of the UK-REACH Study Collaborative Group

**Author notes:** Joint first authors (Christopher A. Martin, Daniel Pan, Joshua Nazareth). Manish Pareek (Chief investigator), Laura Gray (University of Leicester), Laura Nellums (University of Nottingham), Anna L Guyatt (University of Leicester), Catherine Johns (University of Leicester), I Chris McManus (University College London), Katherine Woolf (University College London), Ibrahim Abubakar (University College London), Amit Gupta (Oxford University Hospitals), Keith R Abrams (University of York), Martin D Tobin (University of Leicester), Louise Wain (University of Leicester), Sue Carr (University Hospital Leicester), Edward Dove (University of Edinburgh), Kamlesh Khunti (University of Leicester), David Ford (University of Swansea), Robert Free (University of Leicester). Corresponding author: Dr Manish Pareek, Associate Clinical Professor in Infectious Diseases and Honorary Consultant in Infectious Diseases, Department of Respiratory Sciences, University of Leicester, The corresponding author attests that all listed authors meet authorship criteria and that no others meeting the criteria have been omitted.

## Abstract

**Objectives:** To determine the prevalence and predictors of self-reported access to appropriate personal protective equipment (aPPE) for healthcare workers (HCWs) in the United Kingdom (UK) during the first UK national COVID-19 lockdown (March 2020) and at the time of questionnaire response (December 2020 – February 2021).

**Design:** Two cross sectional analyses using data from a questionnaire-based cohort study.

**Setting:** Nationwide questionnaire from 4^th^ December 2020 to 28^th^ February 2021.

**Participants:** A representative sample of HCWs or ancillary workers in a UK healthcare setting aged 16 or over, registered with one of seven main UK healthcare regulatory bodies.

**Main outcome measure:** Binary measure of self-reported aPPE (access all of the time vs access most of the time or less frequently) at two timepoints: the first national lockdown in the UK (primary analysis) and at the time of questionnaire response (secondary analysis).

**Results:** 10,508 HCWs were included in the primary analysis, and 12,252 in the secondary analysis. 3702 (35.2%) of HCWs reported aPPE at all times in the primary analysis; 6806 (83.9%) reported aPPE at all times in the secondary analysis. After adjustment (for age, sex, ethnicity, migration status, occupation, aerosol generating procedure exposure, work sector, work region, working hours, night shift frequency and trust in employing organisation), older HCWs (per decade increase in age: aOR 1.2, 95% CI 1.16-1.26, p<0.001) and those working in Intensive Care Units (1.61, 1.38 – 1.89, p<0.001) were more likely to report aPPE at all times. Those from Asian ethnic groups compared to White (0.77, 0.67-0.89, p<0.001), those in allied health professional (AHPs) and dental roles (vs those in medical roles; AHPs: 0.77, 0.68 – 0.87, p<0.001; dental: 0.63, 0.49-0.81, p<0.001), and those who saw a higher number of COVID-19 patients compared to those who saw none (≥21 patients 0.74, 0.61-0.90, p=0.003) were less likely to report aPPE at all times in the primary analysis. aPPE at all times was also not uniform across UK regions (reported access being better in South West and North East England than London). Those who trusted their employing organisation to deal with concerns about unsafe clinical practice, compared to those who did not, were twice as likely to report aPPE at all times (2.18, 1.97-2.40, p<0.001). With the exception of occupation, these factors were also significantly associated with aPPE at all times in the secondary analysis.

**Conclusions:** We found that only a third of HCWs in the UK reported aPPE at all times during the period of the first lockdown and that aPPE had improved later in the pandemic. We also identified key sociodemographic and occupational determinants of aPPE during the first UK lockdown, the majority of which have persisted since lockdown was eased. These findings have important public health implications for HCWs, particularly as cases of infection and long-COVID continue to rise in the UK.

**Trial registration:** ISRCTN 11811602

**What is already known on this topic:** Access to personal protective equipment (PPE) is crucial to protect healthcare workers (HCWs) from infection. Limited data exist concerning the prevalence of, and factors relating to, PPE access for HCWs in the United Kingdom (UK) during the COVID-19 pandemic.

**What this study adds:** Only a third of HCWs reported having access to appropriate PPE all of the time during the first UK national lockdown. Older HCWs, those working in Intensive Care Units and those who trusted their employing organisation to deal with concerns about unsafe clinical practice, were more likely to report access to adequate PPE. Those from Asian ethnic groups (compared to White ethnic groups) and those who saw a high number of COVID-19 were less likely to report access to adequate PPE. Our findings have important implications for the mental and physical health of HCWs working during the pandemic in the UK.

## Introduction

As of August 2021, over 6 million people in the United Kingdom (UK) have been infected with Severe Acute Respiratory Syndrome Coronavirus-2 (SARS-CoV-2) leading to substantial morbidity, mortality and demands on health services.[1] Healthcare workers (HCWs) are at significantly higher risk of infection than the general population.[2]

Effective use of personal protective equipment (PPE) might prevent SARS-CoV-2 transmission,[3] however, large numbers of HCWs in the UK have become infected with SARS-CoV-2 whilst working on the frontline. Public Health England estimated that 73% of infections in UK HCWs during the first wave of the COVID-19 pandemic were due to nosocomial transmission;[4] Amnesty International reported in December 2020 that the UK had the second highest rate of COVID-19 related deaths in HCWs in the world.[5] Anecdotal reports exist of limited access to PPE by HCWs and a survey of UK doctors conducted by the British Medical Association during the first wave of the pandemic found that there were self-reported shortages of PPE in both primary and secondary care, however to date, no large studies have examined the issue of PPE availability in detail.[6]

Accordingly, using data from the United Kingdom Research study into Ethnicity And COVID-19 outcomes in Healthcare workers (UK-REACH), we conducted cross sectional analyses of self-reported access to appropriate PPE in a cohort of HCWs at two timepoints during the COVID-19 pandemic in the UK. We sought to determine the occupational and demographic predictors of PPE access, hypothesising that PPE access was not equivalent across HCWs working in the UK and that some HCW groups had more limited access to PPE than others.

## Methods

### Overview

The United Kingdom Research study into Ethnicity And COVID-19 Outcomes in Healthcare workers (UK-REACH), incorporates six studies which aim to establish the impact of the COVID-19 pandemic on UK HCWs, particularly those from ethnic minority groups. This analysis utilises data generated by the baseline questionnaire of the UK-REACH prospective nationwide cohort study. The cohort study has been described in the published study protocol as well as in previous work using the same dataset.[7,8] Details of the measures included in the questionnaire can be found in the data dictionary (https://www.uk-reach.org/data-dictionary).

### Study population

We included National Health Service (NHS) and non-NHS HCWs (including ancillary workers in a healthcare setting) aged 16 years or older and/or registered with one of seven UK professional healthcare regulatory bodies (see supplementary information for a list of participating regulators).

### Recruitment

We have previously described recruitment into the cohort study.[7,8] Briefly, between 4^th^ December 2020 and 28^th^ February 2021, emails with a link to the study website were distributed to HCWs by professional regulators and recruited NHS sites. To take part, eligible HCWs had to visit the website, create a user profile and provide informed consent. The sample was supplemented by recruitment of participants directly through healthcare trusts and advertising on social media / newsletters. We report participation rates as recommended by the Checklist for Reporting Results of Internet E-Surveys (CHERRIES).[9,10]

### Outcome measures

The main outcome measure was access to PPE at two timepoints (see below). We derived a binary measure from a questionnaire item concerning how often a HCW reported access to appropriate PPE with answers on a five point scale (“not at all” through to “all the time”).

For the main analyses we categorised HCWs as either reporting access to appropriate PPE at all times or lacking access to appropriate PPE at least some of the time. We derived a separate binary measure from a different threshold (most of the time or more often vs some of the time or less often) and used this in sensitivity analyses (see supplementary table 1 for the derivation of both measures). We asked participants about PPE access at two timepoints:

1. At the start of the first national lockdown in the UK (23^rd^ March 2020) – used as an outcome measure in the **primary analysis**
2. At the time of answering the questionnaire (December 2020 – February 2021) – used as an outcome measure in the **secondary analysis**

### Covariates

We selected predictor variables that might be associated with the outcome *a priori*, based on existing literature and expert opinion. These are detailed below:

- Demographic characteristics (age, sex, ethnicity [5 categories used by the Office for National Statistics], migration status).[11]
- Occupational factors (job role, area of work, number of confirmed/suspected COVID-19 patients seen per week, exposure to aerosol generating procedures [AGPs], hours worked per week and night shift frequency).
- UK region of workplace.
- Trust in employer to address a concern about unsafe clinical practice – a binary measure derived from a question adapted from the NHS staff survey.[12]

Occupational variables used in the analyses reflect the participants’ occupational circumstances during the first national lockdown in the UK for the primary analysis or at the time they answered the questionnaire for the secondary analysis. Participants could select multiple, non-mutually exclusive areas in which they work, and therefore the work areas variables are coded as ‘dummy’ variables (i.e. all those that selected that area vs all those that did not).

A description of each variable and how it was derived from questionnaire responses can be found in Supplementary Table 2.

### Statistical analysis

Participants with missing outcome data, and those who answered ‘not applicable’ to the question around PPE access were excluded. This was because these HCWs were not likely to require PPE as part of their healthcare role and therefore should not be included in the present analyses. Participants not working during lockdown or at the time of questionnaire response were excluded from the primary and secondary analyses respectively, so that the relevant occupational predictors could be included in the models.

Categorical variables were summarised as count and percentage, and non-normally distributed continuous variables as median and interquartile range (IQR). Logistic regression was used to derive unadjusted and adjusted odds ratios (ORs and aOR) describing the relation between covariates and PPE access.

Multiple imputation was used to replace missing data in all logistic regression models. Rubin’s Rules were used to combine the parameter estimates and standard errors from 10 imputations into a single set of results.[13] To ensure the use of multiple imputation did not significantly affect our results we performed a sensitivity analysis using only complete cases. All analyses were conducted using Stata 17.

### Ethical approval

The study was approved by the Health Research Authority (Brighton and Sussex Research Ethics Committee; ethics reference: 20/HRA/4718). All participants gave informed consent.

### Involvement and engagement

We worked closely with a Professional Expert Panel of HCWs from a range of ethnic backgrounds and occupations as well as with national and local organisations (see study protocol).[7]

## Results

### Recruitment and formation of the cohort

Formation of the cohorts is shown in Figure 1. Cohort recruitment has been described in a previous publication.[8] In brief, between 4^th^ December and 28^th^ February, 1,052,875 emails were sent from regulators. 46% of the emails were received/opened; 26,592 users created a study profile and 17,981 consented to participate. 15,199 HCWs started the questionnaire; 10,508 HCWs were included in the primary analysis (PPE access during lockdown) and 12,252 in the secondary analysis (PPE access at the time of questionnaire response). A summary of missing data for each variable of interest is shown in supplementary table 3.

**Figure 1.**
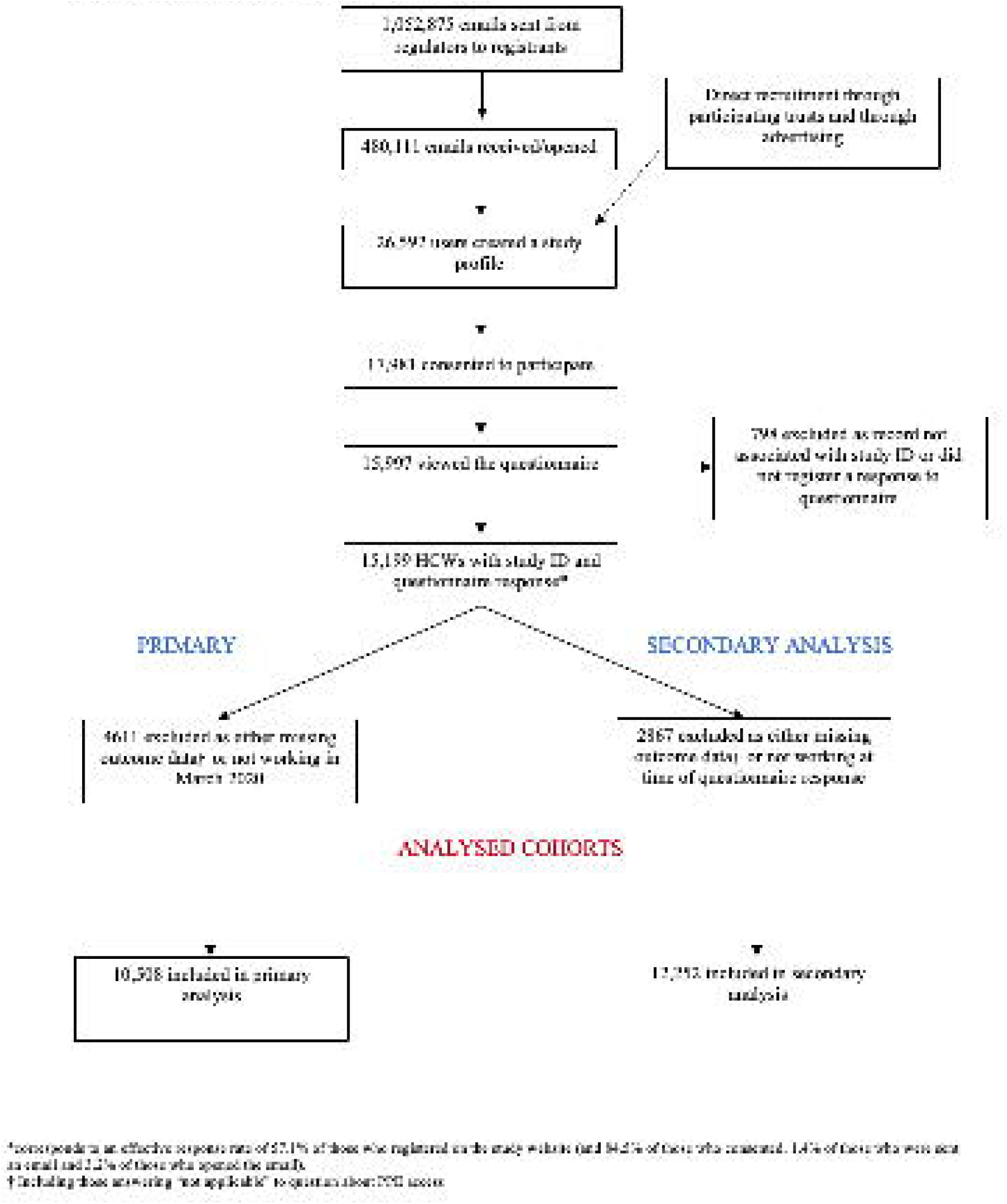
Formation of the analysed cohorts.

### Description of the analysed cohort

Table 1 shows the demographic and occupational characteristics of the 10,508 HCWs who were working during lockdown in the first wave. The median age was 45 (IQR 34 – 54); most respondents were female (74.7%). 30% of participants were from ethnic minority groups (19.9% Asian, 4.4% Black, 4.1% Mixed, 2.1% Other). Description of the 12,252 HCWs working at the time of questionnaire response is also shown in Table 1.

**Table 1.** Description of cohort.

| Variable | Working during lockdown<br>(primary analysis)<br>N=10,508 | Working at the time of<br>questionnaire response<br>(secondary analysis)<br>N=12,252 |
| --- | --- | --- |
| **Age**, med(IQR) | 45 (34 – 54) | 44 (34 – 54) |
| **Sex** |  |  |
| Male | 2653 (25.3%) | 2939 (24.1%) |
| Female | 7829 (74.7%) | 9281 (76.0%) |
| **Ethnicity** |  |  |
| White | 7066 (69.5%) | 8261 (69.7%) |
| Asian | 2026 (19.9%) | 2327 (19.6%) |
| Black | 444 (4.4%) | 511 (4.3%) |
| Mixed | 412 (4.1%) | 491 (4.4%) |
| Other | 218 (2.1%) | 258 (2.2%) |
| **Migration status** |  |  |
| Born in UK | 7517 (72.9%) | 8800 (73.2%) |
| Born abroad | 2801 (27.2%) | 3225 (26.8%) |
| **Occupation** |  |  |
| Doctor or medical support | 2632 (26.0%) | 2788 (23.7%) |
| Nurse, NA or Midwife | 2416 (23.9%) | 2546 (21.6%) |
| Allied Health Professional* | 4232 (41.9%) | 5158 (43.8%) |
| Dental | 382 (3.8%) | 765 (6.5%) |
| Admin, estates or other | 450 (4.5%) | 525 (4.5%) |
| **Work sector** |  |  |
| Non NHS | 1093 (10.9%) | 1849 (15.3%) |
| NHS | 8981 (89.2%) | 10274 (84.8%) |
| **Work areas** |  |  |
| Ambulance | 427 (4.1%) | 485 (4.0%) |
| Emergency Department | 1052 (10.0%) | 1046 (8.6%) |
| Intensive Care Unit | 1014 (9.7%) | 843 (6.9%) |
| Hospital Inpatient | 2960 (28.3%) | 3088 (25.3%) |
| Psychiatric hospital | 325 (3.1%) | 327 (2.7%) |
| Nursing or Care Home | 255 (2.4%) | 299 (2.5%) |
| **Aerosol generating procedure exposure** |  |  |
| Less than weekly exposure | 7777 (76.5%) | 9259 (75.8%) |
| At least weekly exposure | 2396 (23.6%) | 2953 (24.2%) |
| **Night shift pattern** |  |  |
| Never works nights | 6990 (67.3%) | 8629 (73.3%) |
| Works nights less than weekly | 1948 (18.8%) | 1966 (16.7%) |
| Works nights weekly or always | 1445 (13.9%) | 1179 (10.0%) |
| **Number of SARS-CoV-2 positive patients**<br>**attended to per week (with physical**<br>**contact)** |  |  |
| None | 5601 (53.9%) | 8051 (66.7%) |
| 1 – 5 | 2386 (23.0%) | 2362 (19.5%) |
| 6 – 20 | 1645 (15.8%) | 1180 (9.8%) |
| ≥ 21 | 765 (7.4%) | 485 (4.0%) |
| **Work region** |  |  |
| London | 1344 (14.5%) | 1575 (14.5%) |
| South East England | 1219 (13.1%) | 1433 (13.2%) |
| South West England | 826 (8.9%) | 975 (9.0%) |
| East of England | 752 (8.1%) | 919 (8.5%) |
| East Midlands | 1022 (11.0%) | 1176 (10.9%) |
| West Midlands | 804 (8.7%) | 947 (8.7%) |
| North East England | 440 (4.7%) | 482 (4.5%) |
| North West England | 1070 (11.5%) | 1218 (11.2%) |
| Yorkshire and the Humber | 753 (8.1%) | 897 (8.3%) |
| Wales | 318 (3.4%) | 371 (3.4%) |
| Scotland | 609 (6.5%) | 694 (6.4%) |
| Northern Ireland | 134 (1.4%) | 151 (1.4%) |
| **Trust in employer (to address a concern**<br>**about unsafe clinical practice)** |  |  |
| Does not trust employer | 3071 (30.7%) | 3530 (29.5%) |
| Trusts employer | 6945 (69.3%) | 8439 (70.5%) |

\*Also includes pharmacists, healthcare scientists, ambulance workers and those in optical roles.
Occupational factors other than region of workplace relate to work circumstances during the weeks following the first UK national lockdown on March 23<sup>rd</sup> 2020 in the primary analysis and at the time of questionnaire response in the secondary analysis.
IQR – Interquartile range, NHS – national health service, PPE – personal protective equipment, SARS-CoV-2 – severe acute respiratory syndrome coronavirus-2

### Univariable analysis

Table 2 shows demographic and occupational characteristics of HCWs included in the primary analysis, stratified by PPE access and unadjusted odds ratios for the association of these characteristics with reported PPE access. Just over a third (35.2%) of HCWs working during lockdown reported access to appropriate PPE at all times. A significantly smaller proportion of those reporting access to PPE were from Asian ethnic groups than those not reporting access to PPE (16.3% vs 21.9%, OR 0.68, 95%CI 0.61 – 0.76 [reference White]. At the time of questionnaire response (secondary analysis) 83.9% of HCWs reported access to PPE at all times. A description of those included in the secondary analysis stratified by PPE access and unadjusted odds ratios are shown in Supplementary Table 4.

**Table 2.** **Description of those included in the primary analysis by access to personal protective equipment with unadjusted odds ratio**

| Variable | Excluding those not working during lockdown (primary analysis) |  |  |  |
| --- | --- | --- | --- | --- |
|  | Had access to PPE<br>3702 (35.2%) | Did not have access to PPE<br>6806 (64.8%) | Unadjusted OR (95%CI) for access to PPE | P value |
| **Age, med(IQR)** | 47 (38 – 55) | 43 (33 – 53) | 1.25 (1.21 – 1.30) | <0.001 |
| **Sex** |  |  |  |  |
| Male | 915 (24.8%) | 1738 (25.6%) | Ref | - |
| Female | 2782 (75.3%) | 5047 (74.4%) | 1.05 (0.95 – 1.19) | 0.33 |
| **Ethnicity** |  |  |  |  |
| White | 2649 (73.8%) | 4417 (67.2%) | Ref | - |
| Asian | 586 (16.3%) | 1440 (21.9%) | 0.68 (0.61 – 0.76) | <0.001 |
| Black | 145 (4.0%) | 299 (4.6%) | 0.81 (0.66 – 0.99) | 0.04 |
| Mixed | 141 (3.9%) | 271 (4.1%) | 0.88 (0.71 – 1.08) | 0.23 |
| Other | 71 (2.0%) | 147 (2.2%) | 0.80 (0.60 – 1.07) | 0.13 |
| **Migration status** |  |  |  |  |
| Born in UK | 2756 (75.6%) | 4761 (71.4%) | Ref | - |
| Born abroad | 892 (24.5%) | 1909 (68.2%) | 0.81 (0.74 – 0.88) | <0.001 |
| **Occupation** |  |  |  |  |
| Doctor or medical support | 934 (26.2%) | 1698 (25.9%) | Ref | - |
| Nurse, NA or Midwife | 913 (25.6%) | 1503 (23.0%) | 1.12 (1.00 – 1.25) | 0.05 |
| Allied Health Professional* | 1419 (39.8%) | 2813 (43.0%) | 0.93 (0.84 – 1.03) | 0.15 |
| Dental | 118 (3.3%) | 264 (4.0%) | 0.83 (0.66 – 1.04) | 0.11 |
| Admin, estates or other | 181 (5.1%) | 269 (4.1%) | 1.23 (1.00 – 1.51) | 0.05 |
| **Work sector** |  |  |  |  |
| Non NHS | 432 (12.3%) | 661 (10.1%) | Ref | - |
| NHS | 3108 (87.7%) | 5873 (89.9%) | 0.82 (0.72 – 0.93) | 0.002 |
| **Work areas** |  |  |  |  |
| Ambulance | 119 (3.2%) | 308 (4.5%) | 0.70 (0.56 – 0.87) | 0.001 |
| Emergency Department | 311 (8.4%) | 741 (10.9%) | 0.75 (0.65 – 0.86) | <0.001 |
| Intensive Care Unit | 406 (11.0%) | 608 (9.0%) | 1.25 (1.10 – 1.43) | 0.001 |
| Hospital Inpatient | 980 (26.5%) | 1980 (29.2%) | 0.88 (0.80 – 0.96) | 0.004 |
| Psychiatric hospital | 92 (2.5%) | 233 (3.4%) | 0.72 (0.56 – 0.92) | 0.008 |
| Nursing or Care Home | 96 (2.6%) | 159 (2.3%) | 1.12 (0.86 – 1.44) | 0.40 |
| **Aerosol generating procedure exposure** |  |  |  |  |
| Less than weekly exposure | 2822 (76.5%) | 5114 (75.4%) | Ref | - |
| At least weekly exposure | 868 (23.5%) | 1665 (24.6%) | 0.94 (0.86 – 1.04) | 0.23 |
| **Night shift pattern** |  |  |  |  |
| Never works nights | 2520 (68.9%) | 4470 (66.5%) | Ref | - |
| Works nights less than weekly | 685 (18.7%) | 1263 (18.8%) | 0.96 (0.86 – 1.06) | 0.42 |
| Works nights weekly or always | 455 (12.4%) | 990 (14.7%) | 0.82 (0.72 – 0.92) | 0.001 |
| **Hours worked per week, med(IQR)** | 37 (30 – 40) | 38 (30 – 40) | 0.99 (0.99 – 1.00) | <0.001 |
| **Number of SARS-CoV-2 positive patients attended to per week (with physical contact)** |  |  |  |  |
| None | 2135 (58.3%) | 3466 (51.5%) | Ref | - |
| 1 – 5 | 788 (21.5%) | 1598 (23.7%) | 0.80 (0.72 – 0.88) | <0.001 |
| 6 – 20 | 523 (14.3%) | 1122 (16.7%) | 0.75 (0.67 – 0.85) | <0.001 |
| ≥ 21 | 219 (6.0%) | 546 (8.1%) | 0.65 (0.55 – 0.77) | <0.001 |
| **Work region** |  |  |  |  |
| London | 421 (12.7%) | 923 (15.4%) | Ref | - |
| South East England | 421 (12.7%) | 798 (13.3%) | 1.16 (0.98 – 1.38) | 0.27 |
| South West England | 337 (10.2%) | 489 (8.2%) | 1.48 (1.24 – 1.78) | <0.001 |
| East of England | 243 (7.4%) | 509 (8.5%) | 1.06 (0.88 – 1.28) | 0.56 |
| East Midlands | 376 (11.4%) | 646 (10.8%) | 1.25 (1.05 – 1.49) | 0.10 |
| West Midlands | 285 (8.6%) | 519 (8.7%) | 1.19 (0.99 – 1.42) | 0.02 |
| North East England | 183 (5.5%) | 257 (4.3%) | 1.52 (1.22 – 1.89) | 0.02 |
| North West England | 359 (10.9%) | 711 (11.9%) | 1.10 (0.93 – 1.30) | 0.49 |
| Yorkshire and the Humber | 282 (8.5%) | 471 (7.9%) | 1.32 (1.10 – 1.59) | 0.10 |
| Wales | 112 (3.4%) | 206 (3.4%) | 1.20 (0.92 – 1.57) | 0.37 |
| Scotland | 229 (6.9%) | 380 (6.4%) | 1.27 (1.04 – 1.55) | 0.02 |
| Northern Ireland | 59 (1.8%) | 75 (1.3%) | 1.66 (1.16 – 2.38) | 0.006 |
| **Trust in employer (to address a concern about unsafe clinical practice)** |  |  |  |  |
| Does not trust employer | 689 (19.6%) | 2382 (36.6%) | Ref | - |
| Trusts employer | 2820 (80.4%) | 4125 (63.4%) | 2.27 (2.06 – 2.50) | <0.001 |

\*Also includes pharmacists, healthcare scientists, ambulance workers and those in optical roles.
Occupational factors other than region of workplace relate to work circumstances during the weeks following the first UK national lockdown on March 23<sup>rd</sup> 2020
IQR – Interquartile range, NHS – national health service, OR – odds ratio, PPE – personal protective equipment, SARS-CoV-2 – severe acute respiratory syndrome coronavirus-2

### Multivariable analysis

#### Primary analysis: PPE access during lockdown

Table 3 shows adjusted odds ratios for PPE access during the first UK lockdown. On multivariable logistic regression analysis, younger HCWs, as well as those from Asian ethnic groups (compared to those of White ethnicity), allied health professionals and dentists (compared to those in medical roles) were all less likely to report access to PPE at all times during the first lockdown (Figure 2a). Those who had regular physical contact with confirmed or suspected COVID-19 patients were less likely to report access to appropriate PPE at all times compared to those who did not (aOR for PPE access in those who saw 21 or more COVID-19 patients a week compared to those who saw none: 0.74, 95% CI 0.61-0.90). HCWs working in London were less likely to report PPE access at all times compared to South West or North East England, and those who indicated trust in their employer to address concerns about unsafe clinical practice were twice as likely to report PPE access at all times, compared to those who reported the opposite (aOR 2.18, 95% CI 1.97-2.40).

**Table 3.** **Multivariable analysis of factors associated with PPE access at the start of the first UK national lockdown and at the time of answering the questionnaire**

| Variable | Access to PPE during first national lockdown (n=10,508) |  | Access to PPE at the time of response (n=12,252) |  |
| --- | --- | --- | --- | --- |
|  | aOR (95% CI) | p value | aOR (95% CI) | p value |
| **Age\*** | 1.21 (1.16 – 1.26) | <0.001 | 1.18 (1.13 – 1.24) | <0.001 |
| **Sex** |  |  |  |  |
| Male | Ref | - | Ref | - |
| Female | 1.05 (0.94 – 1.17) | 0.37 | 1.10 (0.97 – 1.25) | 0.14 |
| **Ethnicity** |  |  |  |  |
| White | Ref | - | Ref | - |
| Asian | 0.77 (0.67 – 0.89) | <0.001 | 0.79 (0.68 – 0.92) | 0.002 |
| Black | 0.91 (0.73 – 1.14) | 0.41 | 0.83 (0.65 – 1.06) | 0.66 |
| Mixed | 0.96 (0.77 – 1.19) | 0.69 | 0.91 (0.70 – 1.17) | 0.17 |
| Other | 0.86 (0.63 – 1.18) | 0.36 | 0.74 (0.54 – 1.01) | 0.27 |
| **Migration status** |  |  |  |  |
| Born in UK | Ref | - | Ref | - |
| Born abroad | 0.92 (0.82 – 1.04) | 0.18 | 0.71 (0.62 – 0.81) | <0.001 |
| **Occupation** |  |  |  |  |
| Doctor or medical support | Ref | - | Ref | - |
| Nurse, nursing associate or Midwife | 0.89 (0.77 – 1.02) | 0.08 | 1.10 (0.93 – 1.31) | 0.26 |
| Allied health professional<sup>†</sup> | 0.77 (0.68 – 0.87) | <0.001 | 1.27 (1.09 – 1.49) | 0.002 |
| Dental | 0.63 (0.49 – 0.81) | <0.001 | 2.20 (1.57 – 3.08) | <0.001 |
| Admin, estates or other | 0.94 (0.75 – 1.17) | 0.56 | 1.03 (0.77 – 1.38) | 0.86 |
| **Work sector** |  |  |  |  |
| Non NHS | Ref | - | Ref | - |
| NHS | 0.90 (0.78 – 1.03) | 0.13 | 1.04 (0.87 – 1.23) | 0.69 |
| **Work areas** |  |  |  |  |
| Ambulance | 0.96 (0.75 – 1.24) | 0.77 | 0.92 (0.70 – 1.21) | 0.56 |
| Emergency Department | 0.88 (0.76 – 1.03) | 0.12 | 0.84 (0.71 – 1.00) | 0.05 |
| Intensive Care Unit | 1.61 (1.38 – 1.89) | <0.001 | 1.39 (1.12 – 1.71) | 0.002 |
| Hospital Inpatient | 0.99 (0.90 – 1.10) | 0.90 | 0.81 (0.71 – 0.91) | 0.001 |
| Psychiatric hospital | 0.74 (0.57 – 0.95) | 0.02 | 0.77 (0.58 – 1.03) | 0.08 |
| Nursing or Care Home | 1.09 (0.97 – 1.23) | 0.53 | 1.07 (0.74 – 1.53) | 1.07 |
| **Aerosol generating procedure exposure** |  |  |  |  |
| Less than weekly exposure | Ref | - | Ref | - |
| At least weekly exposure | 1.09 (0.97 – 1.23) | 0.14 | 1.14 (0.99 – 1.31) | 0.07 |
| **Night shift pattern** |  |  |  |  |
| Never works nights | Ref | - | Ref | - |
| Works nights less than weekly | 1.19 (1.05 – 1.35) | 0.006 | 0.88 (0.76 – 1.03) | 0.10 |
| Works nights weekly or always | 1.06 (0.91 – 1.23) | 0.42 | 0.81 (0.67 – 0.97) | 0.02 |
| **Hours worked per week** | 0.99 (0.99 – 1.00) | 0.005 | 1.00 (1.00 – 1.01) | 0.56 |
| **Number of SARS-CoV-2 positive patients attended to per week (with physical contact)** |  |  |  |  |
| None | Ref | - | Ref | - |
| 1 – 5 | 0.81 (0.73 – 0.91) | <0.001 | 0.76 (0.66 – 0.87) | <0.001 |
| 6 – 20 | 0.82 (0.72 – 0.95) | 0.009 | 0.69 (0.58 – 0.82) | <0.001 |
| ≥ 21 | 0.74 (0.61 – 0.90) | 0.003 | 0.52 (0.41 – 0.66) | <0.001 |
| **Region of workplace** |  |  |  |  |
| London | Ref | - | Ref | - |
| South East England | 1.06 (0.89 – 1.27) | 0.51 | 1.27 (1.04 – 1.55) | 0.02 |
| South West England or Channel Islands | 1.31 (1.09 – 1.59) | 0.005 | 1.38 (1.10 – 1.74) | 0.006 |
| East of England | 1.00 (0.82 – 1.21) | 0.98 | 1.07 (0.86 – 1.32) | 0.55 |
| East Midlands | 1.15 (0.96 – 1.38) | 0.12 | 1.56 (1.22 – 1.98) | <0.001 |
| West Midlands | 1.17 (0.97 – 1.41) | 0.10 | 1.20 (0.97 – 1.50) | 0.10 |
| North East England | 1.38 (1.09 – 1.74) | 0.006 | 2.19 (1.57 – 3.06) | <0.001 |
| North West England or Isle of Man | 1.04 (0.87 – 1.25) | 0.63 | 1.19 (0.98 – 1.45) | 0.09 |
| Yorkshire and the Humber | 1.19 (0.98 – 1.45) | 0.07 | 1.69 (1.33 – 2.14) | <0.001 |
| Wales | 1.16 (0.88 – 1.53) | 0.29 | 1.30 (0.94 – 1.80) | 0.11 |
| Scotland | 1.21 (0.98 – 1.49) | 0.08 | 1.65 (1.26 – 2.16) | <0.001 |
| Northern Ireland | 1.56 (1.08 – 2.27) | 0.02 | 1.54 (0.92 – 2.59) | 0.10 |
| **Trust in employer (to deal with a concern about unsafe clinical practice)** |  |  |  |  |
| Does not trust employer | Ref | - | Ref | - |
| Trusts employer | 2.18 (1.97 – 2.40) | <0.001 | 2.90 (2.61 – 3.21) | <0.001 |

\*for each decade increase in age. † Also includes pharmacists, healthcare scientists, ambulance workers and those in optical roles. Analyses adjusted for all other variables in the table.
When asked about work areas participants could select multiple answers, therefore the work areas variables are 'dummy' variables comparing all those that did not select an area (reference) with all those that did.
aOR – adjusted odds ratio, NHS – national health service, PPE – personal protective equipment, Ref – reference category for categorical variables, SARS-CoV-2 – severe acute respiratory syndrome coronavirus-2

**Figure 2a:**
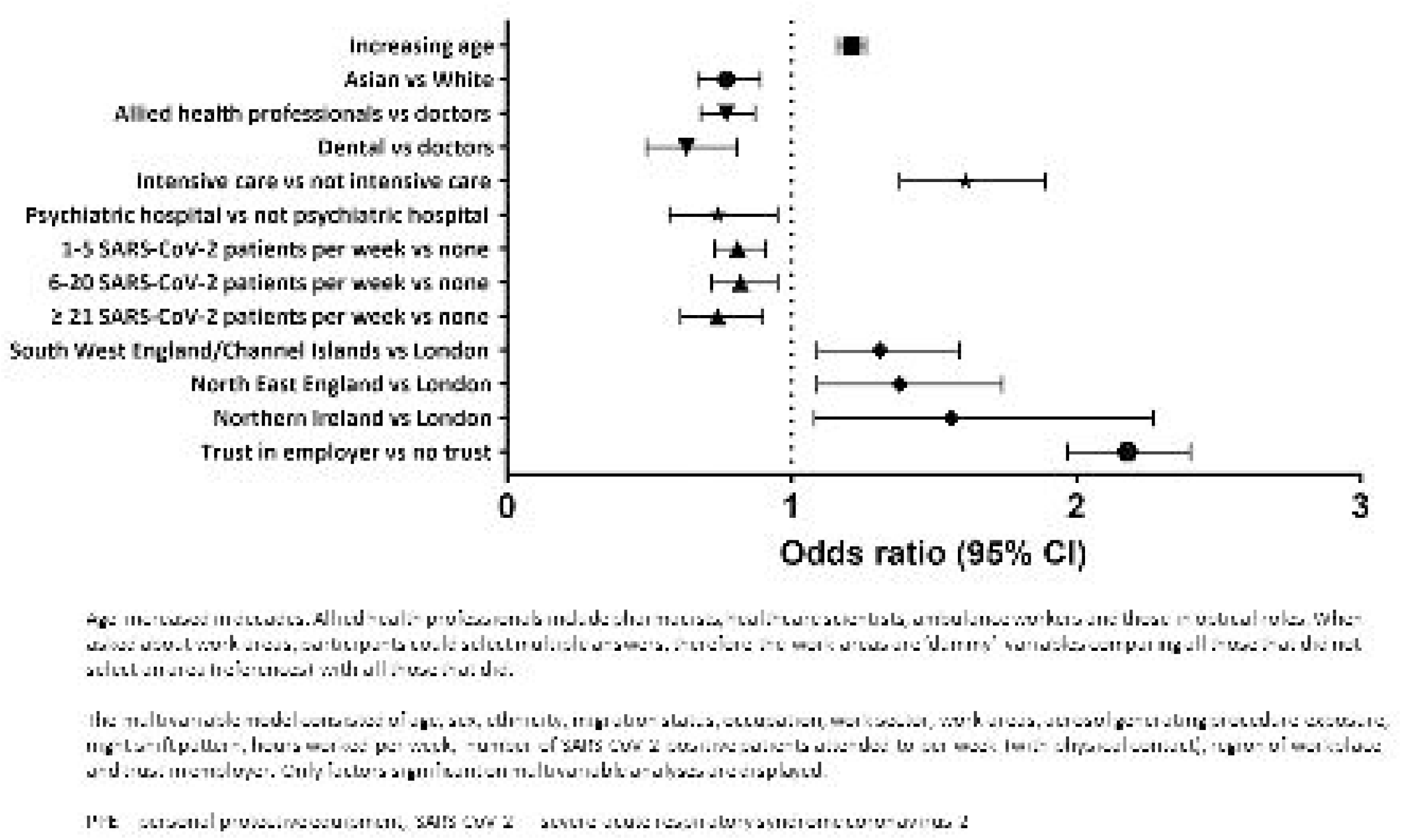
Factors associated with PPE access at the start of the first UK national lockdown on multivariable analysis.

**Figure 2b:**
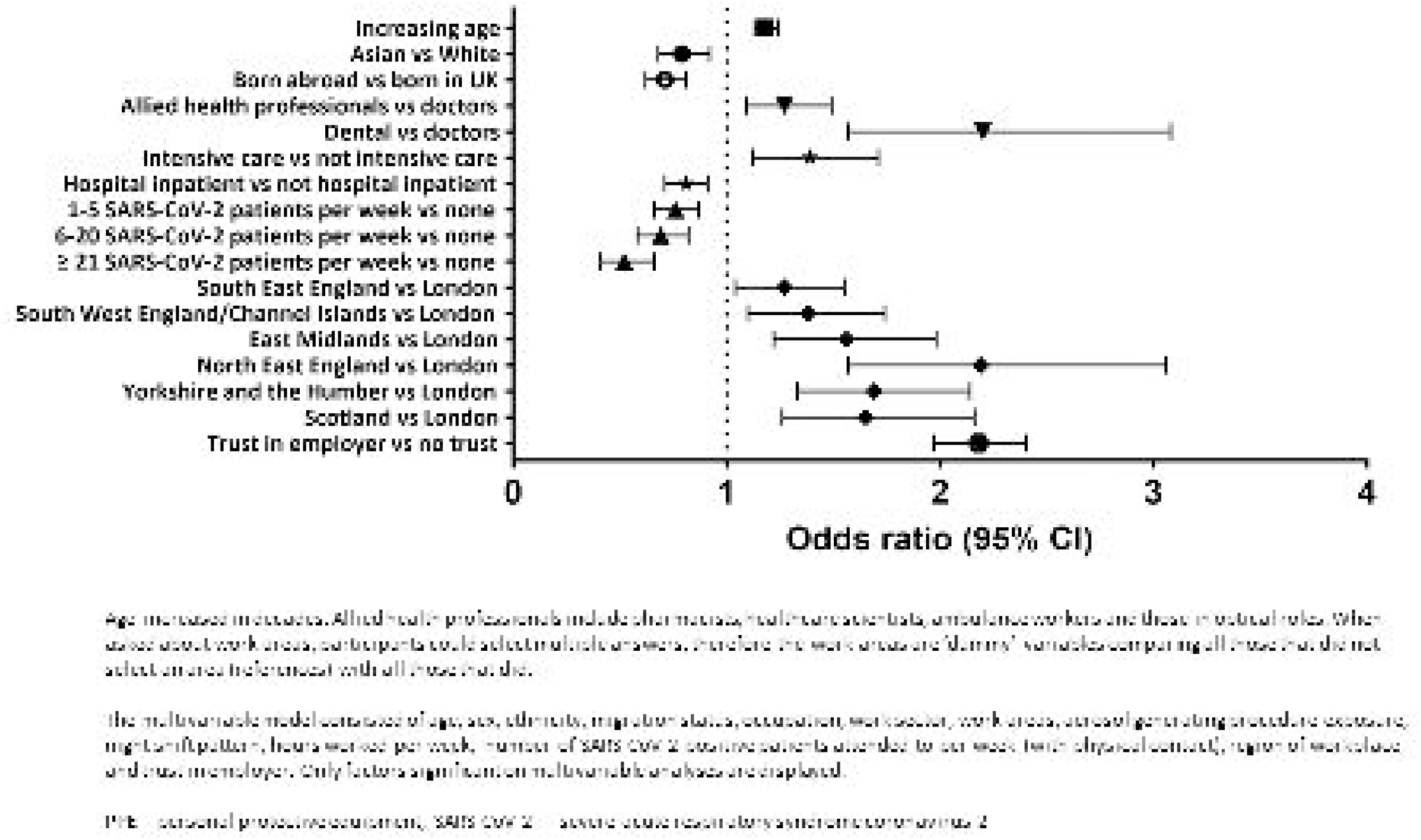
Factors associated with PPE access at the time of answering the questionnaire on multivariable analysis.

#### Secondary analysis: PPE access at time of questionnaire response

Table 3 shows adjusted odds ratios for PPE access at the time of questionnaire response (secondary analysis). Commensurate with findings from the primary analysis, younger HCWs and those from Asian ethnic groups (compared to White groups) were less likely to have access to PPE at all times, with odds ratios similar to those reported in the primary analysis.

By contrast with the primary analysis, HCWs in allied health professional roles and those working in dental roles were more likely to report access to PPE at all times than those in medical roles. The effect of increasing exposure to COVID-19 patients on PPE access was more marked in the secondary analysis, with those attending to ≥21 COVID-19 patients per week being almost half as likely to report access to PPE at all times compared to those that did not attend to any of these patients.

Access to PPE at all times was more likely for those working in South-East England, East and West Midlands, Yorkshire and the Humber and Scotland in addition to South-West and North-East England as compared to those working in London. Trust in employer to address concerns about unsafe clinical practise was a more pronounced independent predictor of PPE access at all times compared to the primary analysis (aOR 2.90, 95%CI 2.61 – 3.21).

### Sensitivity analyses

Changing the threshold of the outcome measure (to access to PPE most of the time or more frequently vs some of the time or less frequently - see Supplementary table 1), did not significantly alter interpretation of the results of the primary analysis (i.e. the majority of significant predictors remain the same). However, differences can be found in the effect of COVID-19 patient exposure (attenuated in the sensitivity analysis compared to the main analysis). Additionally, AGP exposure and working in the NHS increased likelihood of PPE access and all other occupational groups were less likely to report access to PPE than medical staff (see Supplementary table 5). Changing the threshold in the secondary analysis led to only 303 HCWs reporting a lack of access to PPE and thus we considered that the number of events was too low for a multivariable analysis. Repeating the primary analysis using complete cases only did not significantly alter the interpretation of the results (see supplementary table 6).

## Discussion

In this analysis of over 12,000 HCWs across the UK, we found that reported access to appropriate PPE was particularly limited during the first UK lockdown and improved over the course of pandemic. Younger HCWs, Asian ethnic groups (compared to White groups), HCWs who worked in London (compared to multiple regions outside of London), those caring for COVID-19 patients as well as those who reported lack of trust in their employer were less likely to report access to appropriate PPE. HCWs in allied health professional roles were less likely to report access to PPE compared to those in medical roles during lockdown – but this effect reversed by the end of the study period.

Access to appropriate PPE is crucial to preventing HCW infection. When effective PPE is properly donned, removed and discarded, it protects both the HCW who wears it and those with whom the HCW comes into contact. Early in the pandemic, a study in China of 420 doctors and nurses who were deployed to Wuhan for 6 – 8 weeks to care for patients with COVID-19 demonstrated no HCW infection, by both PCR on nasopharyngeal swab and antibody on days 1, 7 and 14 after they had returned; all were fully trained and had access to PPE at all times.[3] Furthermore, a lack of access to PPE for HCWs caring for those with other high-consequence infectious diseases has been shown to be a significant source of physical and mental stress on HCWs as well as their close contacts.[14–16]

Our study provides the first large quantitative summary of reported PPE access amongst HCWs during the COVID-19 pandemic in the literature. We found that only 35% of HCWs in the UK reported having access to adequate PPE at all times during the period of the first lockdown, when at its worst, over 1,000 COVID-19 patients were admitted to hospital a day.[17] Clearly, this finding has implications for HCW infection and SARS-CoV-2 transmission. Our findings are in accordance with qualitative studies that investigated HCWs experiences with PPE during the first wave in the UK. In these studies, HCWs report a major PPE shortage, which in itself was a significant source of anxiety and distress, having a tangible impact on the workforce, resorting to reuse and improvisation of PPE to continue caring for patients when necessary.[18,19] Concern for inadequate PPE stocks may also explain why a higher proportion of allied health professionals and dentists reported lack of PPE access only during the first wave, where they may have been reserved for those looking after hospitalised COVID-19 patients.

We found that the groups which reported limited PPE access were also those that in other studies have been shown to be at highest risk of infection. Indeed, the number of COVID-19 patients seen was an independent negative predictor of PPE access. In a previous study, we found that junior members of staff were more likely to be seropositive for antibodies against SARS-CoV-2 in one UK hospital trust.[20] Junior members of staff are usually younger, and more likely to have more frequent patient contact and fewer administrative and managerial responsibilities, factors that may draw their more senior colleagues away from direct patient care. Furthermore, much of the outpatient work (often undertaken by more senior clinicians) was adapted to include more remote telephone or video consultations during the height of the pandemic, to reduce exposure and therefore the need for PPE. Similarly, HCWs who worked in London hospitals were more likely to have seen a higher number of COVID-19 cases compared to those working in other parts of the country.[21] The more often a HCW sees a patient with COVID-19, the more times one would have had to ‘don’ and ‘doff’ PPE, perpetuating their lack of access if resources during this time were limited. Taken together, these findings add weight to the possibility that lack of PPE access was directly associated with COVID-19 patient contact – which may in part explain the reportedly high number of HCW-associated infections in the UK. It should also be noted that although severe acute COVID-19 might be an unlikely result of SARS-CoV-2 infection amongst younger HCWs, this group may suffer debilitating, prolonged symptoms as a result of ‘long COVID’. Therefore, there may be severe implications both for the individual HCW and the healthcare workforce as a whole (due to absences from work necessitated by the illness).

Of concern, we observed that those from Asian ethnic groups, as well as those who report lack of trust in their employers were less likely to report adequate PPE access compared to White groups or those who reported trust in their employers respectively. Should lack of PPE access be directly related to risk of infection, it may partially explain why HCWs from ethnic minority groups are disproportionately affected by COVID-19.[22–25] Our findings suggest that disparities continue to exist within UK healthcare organisations, the reasons which may be complex.[24] Within the context of HCWs, this could be due to inequities in accessing the right equipment for the tasks required – which in turn can only lead to a further downward spiral of mistrust. It is important that healthcare organisations recognise that such disparities continue to exist and open dialogues with their staff, so that barriers to accessing adequate PPE can be identified and addressed. Furthermore, in light of these findings, it is even more vital that detailed occupational risk assessments that take account of ethnicity are undertaken for UK HCWs.[26]

Our study has limitations. As with any consented observational study, there is potential for self-selection bias. We may be reporting only HCWs’ perspectives regarding what is ‘adequate’ PPE rather than lack of access, since significant variation across a range of different clinical procedures exists for PPE between the UK, other countries and the World Health Organisation.[27] However, the large difference in the proportion of HCWs reporting access to PPE in the primary and secondary analyses provides evidence against this, given that UK PPE guidelines did not change in the intervening time. Our findings relating to trust in employer might indicate reporting bias (i.e. those that did not trust their employer to deal with a concern about unsafe clinical practice might be more likely to report a lack of PPE access in their workplace than those that did). Additionally, given the cross sectional nature of the analyses we cannot determine the direction of causality in this association. Finally, we are asking HCWs to recall their experiences of the last year and thus responses may be prone to recall bias. However, UK-REACH is one of the largest and most comprehensive HCW databases in the world to date, and if only a third of 12,000 HCWs report adequate PPE access, this is difficult for policymakers to ignore. Furthermore, risk factors for lack of PPE access were still present in the secondary analysis, which is less prone to recall bias.

In summary, we have demonstrated that significant proportions of HCWs in the UK reported a lack of access to appropriate PPE over the course of the COVID-19 pandemic and that PPE access was particularly limited during the first national lockdown compared to later in the pandemic. We have also determined key predictors of PPE access. Importantly we show that the demographic and occupational groups who were less likely to report access to PPE overlap with those facing a disproportionate risk of infection. Our study provides evidence of the extraordinary occupational hazard faced by UK frontline HCWs over the course of the pandemic, which has major implications for their physical and mental health, as well as that of their friends and families. Healthcare organisations should urgently implement strategies to understand and address loss of trust from their employees and combat institutional and structural discrimination.

## Data Availability

To access data or samples produced by the UK-REACH study, the working group representative must first submit a request to the Core Management Group by contacting the UK-REACH Project Manager in the first instance. For ancillary studies outside of the core deliverables, the Steering Committee will make final decisions once they have been approved by the Core Management Group. Decisions on granting the access to data/materials will be made within eight weeks.

Third party requests from outside the Project will require explicit approval of the Steering Committee once approved by the Core Management Group.

Note that should there be significant numbers of requests to access data and/or samples then a separate Data Access Committee will be convened to appraise requests in the first instance.

https://www.uk-reach.org/data-dictionary

## Acknowledgements

We would like to thank all the healthcare workers who took part in this study when the NHS was under immense pressure. We wish to acknowledge the members of the UK-REACH Professional Expert Panel (Amir Burney, Association of Pakistani Physicians of Northern Europe; Tiffanie Harrison; London North West University Healthcare NHS Trust; Ahmed Hashim, Sudanese Doctors Association; Sandra Kazembe, University Hospitals Leicester NHS Trust; Susie M. Lagrata (Co-chair), Filipino Nurses Association-UK & University College London Hospitals NHS Foundation Trust; Satheesh Mathew, British Association of Physicians of Indian Origin; Juliette Mutuyimana, Kingston Hospitals NHS Trust; Padmasayee Papineni (Co-chair), London North West University Healthcare NHS Trust), and the Steering and Advisory Group, and SERCO, as well as the following people for their support in setting up the study from the regulatory bodies: Kerrin Clapton and Andrew Ledgard (General Medical Council), Caroline Kenny (Nursing and Midwifery Council), David Teeman and Lisa Bainbridge (General Dental Council), My Phan and John Tse (General Pharmaceutical Council), Angharad Jones and Marcus Dye (General Optical Council), Charlotte Rogers (The Health and Care Professions Council) and Mark Neale (Pharmaceutical Society of Northern Ireland).

We would also like to acknowledge the following trusts and sites who recruited participants to the study: Nottinghamshire Healthcare NHS Foundation Trust, University Hospitals Leicester, Lancashire Teaching Hospitals NHS Foundation Trust, Northumbria Healthcare, Berkshire Healthcare, Derbyshire Healthcare NHS Foundation Trust, South Tees NHS Foundation Trust, Birmingham and Solihull NHS Foundation Trust, Affinity Care, Royal Brompton and Harefield, Sheffield Teaching Hospitals, St George’s Hospital, Yeovil District Hospital, Lewisham and Greenwich NHS Trust, Black Country Community Healthcare NHS Foundation Trust, Sussex Community NHS Foundation Trust, South Central Ambulance Service, University Hospitals Coventry and Warwickshire, University Hospitals Southampton NHS Foundation Trust, London Ambulance Trust, Royal Free, Birmingham Community Healthcare NHS Foundation Trust, Central London Community Healthcare, Chesterfield Royal Hospital, Bridgewater Community Healthcare, Northern Borders, County Durham and Darlington Foundation Trust, Walsall Healthcare NHS Trust.

## References

1 Center for Systems Science and Engineering at Johns Hopkins University. COVID-19 Dashboard. 2021.https://coronavirus.jhu.edu/map.html (accessed 16 Aug 2021).

2 Nguyen LH, Drew DA, Graham MS, et al. Risk of COVID-19 among front-line health-care workers and the general community: a prospective cohort study. Lancet Public Heal Published Online First: 2020. doi:10.1016/s2468-2667(20)30164-x

3 Liu M, Cheng SZ, Xu KW, et al. Use of personal protective equipment against coronavirus disease 2019 by healthcare professionals in Wuhan, China: Cross sectional study. BMJ 2020;369:6–11. doi:10.1136/bmj.m2195

4 Evans S, Agnew E, Vynnycky E, et al. The impact of testing and infection prevention and control strategies on within-hospital transmission dynamics of COVID-19 in English hospitals. Philos Trans R Soc B Biol Sci 2021;376. doi:10.1098/rstb.2020.0268

5 Amnesty International UK. UK among highest COVID-19 health worker deaths in the world. 2020.https://www.amnesty.org.uk/press-releases/uk-among-highest-covid-19-health-worker-deaths-world (accessed 16 Aug 2021).

6 Cooper K. Most doctors still lack protective equipment, finds survey. 2020.https://www.bma.org.uk/news-and-opinion/most-doctors-still-lack-protective-equipment-finds-survey (accessed 8 Sep 2021).

7 Teece L, Gray LJ, Melbourne C, et al. United Kingdom Research study into Ethnicity and COVID-19 outcomes in Healthcare workers (UK-REACH): A retrospective cohort study using linked routinely collected data, study protocol. BMJ Open 2021;11. doi:10.1136/bmjopen-2020-046392

8 Woolf K, McManus IC, Martin CA, et al. Ethnic differences in SARS-CoV-2 vaccine hesitancy in United Kingdom healthcare workers: Results from the UK-REACH prospective nationwide cohort study. Lancet Reg Heal - Eur 2021;000:100180. doi:10.1016/j.lanepe.2021.100180

9 Equator Network. Improving the quality of Web surveys: the Checklist for Reporting Results of Internet E-Surveys (CHERRIES) 2013. 2021.https://www.equator-network.org/reporting-guidelines/improving-the-quality-of-web-surveys-the-checklist-for-reporting-results-of-internet-e-surveys-cherries/ (accessed 21 Jun 2021).

10 Eysenbach G. Improving the quality of web surveys: The Checklist for Reporting Results of Internet E-Surveys (CHERRIES). J Med Internet Res 2004;6:1–6. doi:10.2196/jmir.6.3.e34

11 Office for National Statistics United Kingdom Government. Ethnic group, national identity and religion. 2021.https://www.ons.gov.uk/methodology/classificationsandstandards/measuringequality/ethnicgroupnationalidentityandreligion (

12 NHS care quality commission. National NHS Staff Survey 2020. 2020.

13 Mertens BJA, Banzato E, de Wreede LC. Construction and assessment of prediction rules for binary outcome in the presence of missing predictor data using multiple imputation and cross-validation: Methodological approach and data-based evaluation. Biometrical J 2020;62:724–41. doi:10.1002/bimj.201800289

14 Delamou A, Beavogui AH, Kondé MK, et al. Ebola: Better protection needed for Guinean health-care workers. Lancet 2015;385:503–4. doi:10.1016/S0140-6736(15)60193-3

15 Pathmanathan I, O’Connor KA, Adams ML, et al. Rapid assessment of Ebola infection prevention and control needs--six districts, Sierra Leone, October 2014. MMWR Morb Mortal Wkly Rep 2014;63:1172–4. http://www.ncbi.nlm.nih.gov/pubmed/25503922%0Ahttp://www.pubmedcentral.nih.gov/articlerender.fcgi?artid=PMC4584542

16 The Lancet. Ebola: Protection of health-care workers. Lancet 2014;384:2174. doi:10.1016/S0140-6736(14)62413-2

17 UK Government. Healthcare in the UK - Coronavirus in the UK. 2021.https://coronavirus.data.gov.uk/details/healthcare (accessed 16 Aug 2021).

18 Hoernke K, Djellouli N, Andrews L, et al. Frontline healthcare workers’ experiences with personal protective equipment during the COVID-19 pandemic in the UK: A rapid qualitative appraisal. BMJ Open 2021;11. doi:10.1136/bmjopen-2020-046199

19 Vindrola-Padros C, Andrews L, Dowrick A, et al. Perceptions and experiences of healthcare workers during the COVID-19 pandemic in the UK. BMJ Open 2020;10:1–8. doi:10.1136/bmjopen-2020-040503

20 Martin CA, Patel P, Goss C, et al. Demographic and occupational determinants of anti-SARS-CoV-2 IgG seropositivity in hospital staff. J Public Health (Bangkok) 2020;:1–12. doi:10.1093/pubmed/fdaa199

21 Treibel TA, Manisty C, Burton M, et al. COVID-19: PCR screening of asymptomatic health-care workers at London hospital. Lancet 2020;395:1608–10. doi:10.1016/S0140-6736(20)31100-4

22 Pareek M, Bangash M, Pareek N, et al. Ethnicity and COVID-19: an urgent public health research priority. Lancet 2020.

23 Pan D, Martin C, Nazareth J, et al. Ethnic disparities in COVID-19: increased risk of infection or severe disease? Lancet 2021;Accepted,.

24 Pan D, Sze S, Minhas JS, et al. The impact of ethnicity on clinical outcomes in COVID-19: A systematic review. EClinicalMedicine 2020;23:100404. doi:10.1016/j.eclinm.2020.100404

25 Martin CA, Jenkins DR, Minhas JS, et al. Socio-demographic heterogeneity in the prevalence of COVID-19 during lockdown is associated with ethnicity and household size: Results from an observational cohort study. EClinicalMedicine 2020;25:100466. doi:10.1016/j.eclinm.2020.100466

26 Khunti K, Griffiths A, Majeed A, et al. Assessing risk for healthcare workers during the covid-19 pandemic. BMJ 2021;372:1–4. doi:10.1136/bmj.n602

27 Mudalige NL, Sze S, Oyefeso O, et al. To PPE or not to PPE? Making sense of conflicting international recommendations for PPE during chest compressions in patients with COVID-19. Resuscitation 2020;156:146–8. doi:10.1016/j.resuscitation.2020.09.019

