## Supplementary information for "Access to personal protective equipment in healthcare workers during the COVID-19 pandemic in the United Kingdom: results from a nationwide cohort study (UK-REACH)"

Corresponding author: Dr Manish Pareek

**Supplementary information - list of participating healthcare regulators**

- The General Medical Council (GMC)
- The Nursing and Midwifery Council (NMC)
- The General Dental Council (GDC)
- The Health and Care Professions Council (HCPC)
- The General Optical Council (GOC)
- The General Pharmaceutical Council (GPC)
- The Pharmaceutical Society of Northern Ireland (PSNI)

**Supplementary Table 1. Derivation of binary outcome variables**

| Do you (or did you) have access to appropriate personal protective equipment (PPE) at work?* | OUTCOME  MAIN ANALYSES | OUTCOME  SENSITIVITY ANALYSIS |
| --- | --- | --- |
| All the time | Had access | Had access |
| Most of the time | Did not have access | Had access |
| Some of the time | Did not have access | Did not have access |
| Rarely | Did not have access | Did not have access |
| Not at all | Did not have access | Did not have access |
| Not applicable | Excluded | Excluded |

Participants were asked this question with respect to access to PPE at the time of answering the questionnaire and at the start of the first UK national lockdown (starting 23^rd^ March 2020). The available answer options are shown in the white boxes. Participants giving answers corresponding to green boxes were coded as ‘1’ or ‘has access to PPE’, and those giving answers corresponding to red boxes represent were coded as ‘0’ or ‘does not have access to PPE’. Those answering ‘not applicable’ were excluded from the analysis (blue boxes).

**Supplementary table 2. Derivation of covariates from questionnaire data**

| **Variable** | **Description** |
| --- | --- |
| **Age** | Continuous variable. Age in years. Derived from date of birth entered by participants at registration. |
| **Sex** | Binary variable. Participants were asked their sex assigned at birth. |
| **Ethnicity** | Categorical variable. Participants were asked to select their ethnicity from a list of the 18 Office for National Statistics categories:  Asian/Asian British – Indian  Asian/Asian British – Pakistani  Asian/Asian British – Bangladeshi  Asian/Asian British – Chinese  Asian/Asian British - Any other Asian background  Black/African/Caribbean/Black British - African  Black/African/Caribbean/Black British – Caribbean  Black/African/Caribbean/Black British - Any other Black/African/Caribbean background Mixed/Multiple ethnic groups - White and Black Caribbean  Mixed/Multiple ethnic groups - White and Black African  Mixed/Multiple ethnic groups - White and Asian  Mixed/Multiple ethnic groups - Any other Mixed/multiple ethnic background  White - English/Welsh/Scottish/Northern Irish/British  White – Irish  White - Gypsy or Irish Traveller  White - Any other white background  Other ethnic group – Arab  Other ethnic group - Any other ethnic background  These were categorised into the 5 broader Office for National Statistics ethnicity categories (Asian, Black, Mixed, White, Other). |
| **Migration status** | Binary variable. Participants were asked whether they were born in the UK. |
| **Occupation** | Categorical variable. Participants were asked to select their main job/role. Categorised as below:  **Doctor or medical support** - Doctor, Advanced Critical Care Practitioner, Anaesthesia associate, Surgical Care Practitioner, Other medical associate  **Nurse, NA or Midwife -**  Advanced Nurse Practitioner, Healthcare assistant, Maternity support worker, Midwife, Nurse, Nursing Associate, Other nursing and midwifery role,  **Allied Health Professional (including pharmacists, ambulance workers and those in optical roles)** - Arts therapist, Biomedical scientist, Chiropodist/Podiatrist, Clinical scientist, Dietician, Hearing aid dispenser, Occupational therapist, Operating department practitioner, Orthoptist, Physiotherapist, Practitioner psychologist, Prosthetist / Orthotist, Radiographer, Speech and language therapist, Other Allied Health Professional role, Emergency medical , Paramedic , Other ambulance role, OT Support , Phlebotomist, Physiotherapy Assistant, Radiography Other clinical support role , Pharmacist , Pharmacy technician, Other pharmacy role, Optical - Dispensing optician, Optometrist, Other Optical role  **Dental -**  Clinical dental technician, Dental Hygienist, Dental nurse, Dental technician, Dentist, Other dental role  **Admin, estates or other –** Administration, Catering services, Domestic services, Estates services, Porter, Other |
| **Work sector*** | Binary variable (does not work for NHS vs does work for NHS). Participants were asked to select which from the following list of sectors: NHS, Other public sector (e.g. local or national government), Private sector, Private facility temporarily used by the NHS, University / higher education. These were categorised into NHS and non-NHS. |
| **Work areas*** | Binary (dummy) variables. Participants were asked to select from a list of non-mutually exclusive clinical and non-clinical areas. |
| **Aerosol generating procedure exposure*** | Binary variable. Less than weekly vs weekly or more often). Derived from a question that asked how often participants were in a room where aerosol generating procedures are performed. On the following scale: Not applicable, Never, Once a month or less, A few times a month, Once a week, A few times a week, Every day. |
| **Night shift pattern*** | Categorical variable. Participants were asked how often they work night shifts. On the following scale: Not applicable, Never, Less than once a month, Once a month or more, but not every week, Once a week or more, but not every shift, I always work nights. This was categorised as never, less than weekly or weekly/always |
| **Work hours*** | Continuous variable. Participants were asked to free-type how many hours they work per week |
| **Number of SARS-CoV-2 positive patients attended to per week (with physical contact)*** | Ordinal variable. Participants were asked to select how many suspected or confirmed COVID-19 patients they attended to (with physical contact) in a week on the following scale: 0, 1-5, 6-20, 21-50, 51+. This was categorised as 0, 1-5, 6 – 20, ≥21. |
| **Work region** | Categorical variable. Participants were asked to enter the first part of the postcode of their place of work. This was mapped to one of 12 UK regions. |
| **Trust in employer (to address a concern about unsafe clinical practice)** | Binary variable (trusts employer vs does not trust employer). Participants were asked to indicate how much they agree with the following statement “I am confident that my organisation would address my concern [about unsafe clinical practice]”. Participants who agreed or strongly agreed with this statement were coded as trusting their employer. |

*participants were asked the questions in relation to circumstances both during lockdown and at the time of questionnaire response.

**Supplementary table 3. Summary of missing data for primary and secondary analyses**

| **Variable** | **PRIMARY ANALYSIS**  **Missing n(%)** | **SECONDARY ANALYSIS**  **Missing n(%)** |
| --- | --- | --- |
| **Age** | 53 (0.5%) | 66 (0.5%) |
| **Sex** | 26 (0.3%) | 32 (0.3%) |
| **Ethnicity** | 342 (3.3%) | 404 (3.3%) |
| **Migration status** | 190 (1.8%) | 227 (1.9%) |
| **Occupation** | 396 (3.8%) | 470 (3.8%) |
| **Work sector** | 434 (4.1%) | 129 (1.1%) |
| **Work areas**  Ambulance  Emergency Department  Intensive Care Unit  Hospital inpatients  Psychiatric hospital  Nursing or Care Home | 31 (0.3%)  29 (0.3%)  32 (0.3%)  33 (0.3%)  31 (0.3%)  32 (0.3%) | 58 (0.5%)  58 (0.5%)  61 (0.5%)  62 (0.5%)  56 (0.5%)  59 (0.5%) |
| **Aerosol generating procedure exposure** | 39 (0.4%) | 40 (0.3%) |
| **Night shift pattern** | 125 (1.2%) | 478 (3.9%) |
| **Hours worked per week** | 104 (1.0%) | 154 (1.3%) |
| **Number of SARS-CoV-2 positive patients attended to per week (with physical contact)** | 111 (1.1%) | 174 (1.4%) |
| **Region of workplace** | 1217 (11.6%) | 1414 (11.5%) |
| **Trust in employer** | 492 (4.7%) | 283 (2.3%) |

**Supplementary Table 4. Description of those included in the secondary analysis by access to personal protective equipment**

| **Variable** | **Excluding those not working at the start of the first national lockdown**  **(secondary analysis)** | | | |
| --- | --- | --- | --- | --- |
|  | **Had access to PPE**  **10,276 (83.9%)** | **Did not have access to PPE**  **1976 (16.1%)** | **Unadjusted OR (95%CI) for access to PPE** | **P value** |
| **Age**, med(IQR) | 45 (35 – 54) | 40 (31 – 51) | 1.26 (1.21 – 1.31) | <0.001 |
| **Sex**  Male  Female | 2373 (23.1%)  7881 (76.9%) | 566 (28.8%)  1400 (71.2%) | Ref  1.34 (1.21 – 1.50) | -  <0.001 |
| **Ethnicity**  White  Asian  Black  Mixed  Other | 7172 (72.1%)  1785 (18.0%)  396 (4.0%)  404 (4.1%)  189 (1.9%) | 1089 (57.3%)  542 (28.5%)  115 (6.1%)  87 (4.6%)  69 (3.6%) | Ref  0.50 (0.45 – 0.56)  0.53 (0.43 – 0.66)  0.70 (0.55 – 0.89)  0.42 (0.32 – 0.56) | -  <0.001  <0.001  0.003  <0.001 |
| **Migration status**  Born in UK  Born abroad | 7616 (75.4%)  2483 (24.6%) | 1184 (61.5%)  742 (38.5%) | Ref  0.52 (0.47 – 0.58) | -  <0.001 |
| **Occupation**  Doctor or medical support  Nurse, NA or Midwife  Allied Health Professional*  Dental  Admin, estates or other | 2153 (21.7%)  2147 (21.7%)  4440 (44.8%)  712 (7.2%)  457 (4.6%) | 635 (33.9%)  399 (21.3%)  718 (38.3%)  53 (2.8%)  68 (3.6%) | Ref  1.60 (1.39 – 1.84)  1.82 (1.62 – 2.05)  3.98 (2.96 – 5.34)  1.99 (1.53 – 2.60) | -  <0.001  <0.001  <0.001  <0.001 |
| **Work sector**  Non NHS  NHS | 1633 (16.1%)  8534 (83.9%) | 216 (11.0%)  1740 (89.0%) | Ref  0.64 (0.55 – 0.75) | -  <0.001 |
| **Work areas**  Ambulance  Emergency Department  Intensive Care Unit  Hospital inpatient  Psychiatric hospital  Nursing or Care Home | 379 (3.7%)  751 (7.3%)  682 (6.7%)  2373 (23.2%)  257 (2.5%)  259 (2.5%) | 106 (5.4%)  295 (15.0%)  161 (8.2%)  715 (36.4%)  70 (3.6%)  40 (2%) | 0.68 (0.54 – 0.84)  0.45 (0.39 – 0.52)  0.80 (0.67 – 0.96)  0.53 (0.48 – 0.58)  0.70 (0.53 – 0.91)  1.24 (0.89 – 1.75) | 0.001  <0.001  0.02  <0.001  0.009  0.20 |
| **Aerosol generating procedure exposure**  Less than weekly exposure  At least weekly exposure | 7819 (76.3%)  2426 (23.7%) | 1440 (73.2%)  527 (26.8%) | Ref  0.85 (0.76 – 0.94) | -  0.03 |
| **Night shift pattern**  Never works nights  Works nights less than weekly  Works nights weekly or always | 7480 (75.7%)  1506 (15.2%)  897 (9.1%) | 1149 (60.8%)  460 (24.3%)  282 (14.9%) | Ref  0.51 (0.45 – 0.57)  0.49 (0.42 – 0.57) | -  <0.001  <0.001 |
| **Hours worked per week,** med(IQR) | 37 (30 – 40) | 38 (32 – 40) | 0.98 (0.97 – 0.98) | <0.001 |
| **Number of SARS-CoV-2 positive patients attended to per week (with physical contact)**  None  1 – 5  6 – 20  ≥ 21 | 7053 (69.6%)  1860 (18.4%)  882 (8.7%)  339 (3.4%) | 998 (51.3%)  502 (25.8%)  298 (15.3%)  146 (7.5%) | Ref  0.80 (0.72 – 0.88)  0.75 (0.67 – 0.85)  0.65 (0.55 – 0.77) | -  <0.001  <0.001  <0.001 |
| **Work region**  London  South East England  South West England  East of England  East Midlands  West Midlands  North East England  North West England  Yorkshire and the Humber  Wales  Scotland  Northern Ireland | 1243 (13.6%)  1219 (13.3%)  845 (9.2%)  749 (8.2%)  1028 (11.2%)  782 (8.6%)  439 (4.8%)  998 (10.9%)  792 (8.7%)  312 (3.4%)  608 (6.7%)  132 (1.4%) | 332 (19.6%)  214 (12.7%)  130 (7.7%)  170 (10.1%)  148 (8.8%)  165 (9.8%)  43 (2.5%)  220 (13.0%)  105 (6.2%)  59 (3.5%)  86 (5.1%)  19 (1.1%) | Ref  1.16 (0.98 – 1.38)  1.48 (1.24 – 1.78)  1.06 (0.88 – 1.28)  1.25 (1.05 – 1.49)  1.19 (0.99 – 1.42)  1.52 (1.22 – 1.89)  1.10 (0.93 – 1.30)  1.32 (1.10 – 1.59)  1.20 (0.92 – 1.57)  1.27 (1.04 – 1.55)  1.66 (1.16 – 2.38) | -  0.27  <0.001  0.56  0.10  0.02  0.02  0.49  0.10  0.37  0.02  0.006 |
| **Trust in employer (to address a concern about unsafe clinical practice)**  Does not trust employer  Trusts employer | 2512 (25.0%)  7521 (75.0%) | 1018 (52.6%)  918 (47.4%) | Ref  2.27 (2.06 – 2.50) | -  <0.001 |

*for each decade increase in age. † Also includes pharmacists, healthcare scientists, ambulance workers and those in optical roles.

When asked about work areas participants could select multiple answers , therefore the work areas variables are ‘dummy’ variables comparing all those that did not select an area (reference) with all those that did.

NHS – national health service, OR – odds ratio, PPE – personal protective equipment, Ref – reference category for categorical variables, SARS-CoV-2 – severe acute respiratory syndrome coronavirus-2

**Supplementary table 5. Multivariable analysis of factors associated with PPE access during the first UK national lockdown with altered outcome threshold (sensitivity analysis)**

|  | **Access to PPE during first national lockdown**  **(n=10,508)** | |
| --- | --- | --- |
| **Variable** | **aOR (95% CI)** | **p value** |
| **Age*** | 1.13 (1.08 – 1.17) | <0.001 |
| **Sex**  Male  Female | Ref  0.97 (0.88 – 1.08) | -  0.59 |
| **Ethnicity**  White  Asian  Black  Mixed  Other | Ref  0.75 (0.66 – 0.85)  0.78 (0.63 – 0.97)  0.93 (0.75 – 1.15)  0.76 (0.56 – 1.04) | -  <0.001  0.02  0.49  0.08 |
| **Migration status**  Born in UK  Born abroad | Ref  0.95 (0.85 – 1.06) | -  0.38 |
| **Occupation**  Doctor or medical support  Nurse, nursing associate or Midwife  Allied health professional^†^  Dental  Admin, estates or other | Ref  0.82 (0.71 – 0.94)  0.68 (0.60 – 0.77)  0.47 (0.37 – 0.60)  0.64 (0.51 – 0.80) | -  0.004  <0.001  <0.001  <0.001 |
| **Work sector**  Non NHS  NHS | Ref  1.22 (1.06 – 1.40) | -  0.004 |
| **Work areas**  Ambulance  Emergency Department  Intensive Care Unit  Hospital Inpatient  Psychiatric hospital  Nursing or Care Home | 1.20 (0.95 – 1.52)  1.00 (0.86 – 1.16)  2.03 (1.71 – 2.41)  1.15 (1.04 – 1.29)  0.77 (0.61 – 0.98)  1.24 (0.94 – 1.63) | 0.77  0.98  <0.001  0.006  0.03  0.13 |
| **Aerosol generating procedure exposure**  Less than weekly exposure  At least weekly exposure | Ref  1.22 (1.09 – 1.38) | -  0.14 |
| **Night shift pattern**  Never works nights  Works nights less than weekly  Works nights weekly or always | Ref  1.21 (1.06 – 1.37)  1.03 (0.89 – 1.19) | -  0.003  0.69 |
| **Hours worked per week** | 0.99 (0.99 – 1.00) | 0.014 |
| **Number of SARS-CoV-2 positive patients attended to per week (with physical contact)**  None  1 – 5  6 – 20  ≥ 21 | Ref  1.09 (0.97 – 1.22)  1.08 (0.94 – 1.24)  0.79 (0.66 – 0.95) | -  0.15  0.31  0.01 |
| **Region of workplace**  London  South East England  South West England or Channel Islands  East of England  East Midlands  West Midlands  North East England  North West England or Isle of Man  Yorkshire and the Humber  Wales  Scotland  Northern Ireland | Ref  1.06 (0.89 – 1.25)  1.47 (1.21 – 1.78)  1.08 (0.90 – 1.29)  1.21 (1.02 – 1.44)  1.14 (0.95 – 1.37)  1.33 (1.06 – 1.68)  0.99 (0.83 – 1.17)  1.17 (0.97 – 1.42)  1.13 (0.88 – 1.46)  1.13 (0.92 – 1.39)  1.37 (0.93 – 2.01) | -  0.52  <0.001  0.43  0.03  0.16  0.01  0.87  0.11  0.34  0.24  0.11 |
| **Trust in employer** (to deal with a concern about unsafe clinical practice)  Does not trust employer  Trusts employer | Ref  2.34 (2.14 – 2.56) | -  <0.001 |

*for each decade increase in age. † Also includes pharmacists, healthcare scientists, ambulance workers and those in optical roles.

Analyses adjusted for all other variables in the table.

When asked about work areas participants could select multiple answers , therefore the work areas variables are ‘dummy’ variables comparing all those that did not select an area (reference) with all those that did.

aOR – adjusted odds ratio (adjusted for all variables in the table), NHS – national health service, PPE – personal protective equipment, Ref – reference category for categorical variables, SARS-CoV-2 – severe acute respiratory syndrome coronavirus-2

**Supplementary Table 6. Multivariable analysis of factors associated with PPE access during the first UK national lockdown and at the time of questionnaire response including only complete cases (sensitivity analysis)**

|  | **Access to PPE during first national lockdown**  **(n=7819)** | | **Access to PPE at the time of response**  **(n=9377)** | |
| --- | --- | --- | --- | --- |
| **Variable** | **aOR (95% CI)** | **p value** | **aOR (95% CI)** | **p value** |
| **Age*** | 1.22 (1.17 – 1.28) | <0.001 | 1.19 (1.13 – 1.27) | <0.001 |
| **Sex**  Male  Female | Ref  1.06 (0.94 – 1.19) | -  0.37 | Ref  1.16 (1.00 – 1.34) | -  0.05 |
| **Ethnicity**  White  Asian  Black  Mixed  Other | Ref  0.81 (0.69 – 0.94)  0.90 (0.69 – 1.18)  0.92 (0.72 – 1.18)  0.93 (0.64 – 1.34) | -  0.007  0.46  0.50  0.69 | Ref  0.81 (0.69 – 0.97)  0.78 (0.59 – 1.03)  0.89 (0.67 – 1.18)  0.71 (0.49 – 1.03) | -  0.02  0.08  0.43  0.07 |
| **Migration status**  Born in UK  Born abroad | Ref  0.95 (0.83 – 1.09) | -  0.47 | Ref  0.72 (0.62 – 0.84) | -  <0.001 |
| **Occupation**  Doctor or medical support  Nurse, nursing associate or Midwife  Allied health professional^†^  Dental  Admin, estates or other | Ref  0.82 (0.70 – 0.95)  0.71 (0.62 – 0.82)  0.50 (0.37 – 0.68)  0.88 (0.68 – 1.13) | -  0.01  <0.001  <0.001  0.31 | Ref  0.96 (0.79 – 1.17)  1.18 (0.99 – 1.40)  1.73 (1.17 – 2.55)  0.95 (0.68 – 1.33) | -  0.26  0.002  0.006  0.78 |
| **Work sector**  Non NHS  NHS | Ref  1.01 (0.86 – 1.20) | -  0.89 | Ref  1.01 (0.82 – 1.24) | -  0.93 |
| **Work areas**  Ambulance  Emergency Department  Intensive Care Unit  Hospital Inpatient  Psychiatric hospital  Nursing or Care Home | 1.00 (0.75 – 1.34)  0.94 (0.79 – 1.13)  1.71 (1.43 – 2.06)  0.96 (0.86 – 1.09)  0.69 (0.51 – 0.92)  1.16 (0.82 – 1.63) | 0.97  0.50  <0.001  0.56  0.01  0.40 | 0.90 (0.65 – 1.24)  0.79 (0.65 – 0.97)  1.51 (1.18 – 1.93)  0.74 (0.64 – 0.86)  0.72 (0.52 – 0.99)  0.83 (0.55 – 1.24) | 0.52  0.02  0.001  <0.001  0.04  0.37 |
| **Aerosol generating procedure exposure**  Less than weekly exposure  At least weekly exposure | Ref  1.10 (0.95 – 1.26) | -  0.19 | Ref  1.12 (0.95 – 1.32) | -  0.17 |
| **Night shift pattern**  Never works nights  Works nights less than weekly  Works nights weekly or always | Ref  1.15 (0.99 – 1.32)  1.07 (0.90 – 1.27) | -  0.06  0.45 | Ref  0.87 (0.73 – 1.03)  0.91 (0.73 – 1.13) | -  0.11  0.41 |
| **Hours worked per week** | 0.99 (0.99 – 1.00) | 0.001 | 1.00 (0.99 – 1.00) | 0.53 |
| **Number of SARS-CoV-2 positive patients attended to per week (with physical contact)**  None  1 – 5  6 – 20  ≥ 21 | Ref  0.83 (0.73 – 0.95)  0.81 (0.69 – 0.96)  0.75 (0.59 – 0.94) | -  0.006  0.01  0.01 | Ref  0.80 (0.68 – 0.94)  0.71 (0.57 – 0.87)  0.55 (0.41 – 0.73) | -  0.007  0.001  <0.001 |
| **Region of workplace**  London  South East England  South West England or Channel Islands  East of England  East Midlands  West Midlands  North East England  North West England or Isle of Man  Yorkshire and the Humber  Wales  Scotland  Northern Ireland | Ref  1.11 (0.92 – 1.34)  1.32 (1.07 – 1.62)  1.01 (0.81 – 1.25)  1.18 (0.97 – 1.43)  1.28 (1.04 – 1.58)  1.37 (1.06 – 1.76)  1.07 (0.88 – 1.30)  1.19 (0.96 – 1.47)  1.14 (0.86 – 1.53)  1.37 (1.09 – 1.72)  1.95 (1.29 – 2.95) | -  0.27  0.008  0.93  0.10  0.02  0.02  0.49  0.10  0.37  0.007  0.001 | Ref  1.44 (1.15 – 1.79)  1.43 (1.11 – 1.85)  1.02 (0.81 – 1.29)  1.64 (1.29 – 2.09)  1.19 (0.94 – 1.51)  2.40 (1.65 – 3.50)  1.28 (1.02 – 1.59)  1.70 (1.30 – 2.22)  1.22 (0.87 – 1.71)  1.80 (1.34 – 2.40)  1.56 (0.89 – 2.74) | -  0.001  0.005  0.88  <0.001  0.14  <0.001  0.03  <0.001  0.24  <0.001  0.12 |
| **Trust in employer** (to deal with a concern about unsafe clinical practice)  Does not trust employer  Trusts employer | Ref  2.46 (2.19 – 2.76) | -  <0.001 | Ref  3.08 (2.73 – 3.48) | -  <0.001 |

*for each decade increase in age. † Also includes pharmacists, healthcare scientists, ambulance workers and those in optical roles.

Analyses adjusted for all other variables in the table.

When asked about work areas participants could select multiple answers , therefore the work areas variables are ‘dummy’ variables comparing all those that did not select an area (reference) with all those that did.

aOR – adjusted odds ratio (adjusted for all variables in the table), NHS – national health service, PPE – personal protective equipment, Ref – reference category for categorical variables, SARS-CoV-2 – severe acute respiratory syndrome coronavirus-2
